# Why is patient safety a challenge? Insights from the Professionalism Opinions of Medical Students’ (PoMS) Research

**DOI:** 10.1101/2021.07.20.21260739

**Authors:** P McGurgan, K. Calvert, K. Narula, E. Nathan, A. Celenza, C. Jorm

**Affiliations:** UWA School of Medicine, Perth, WA; King Edward Memorial Hospital, PGME, Perth, WA; Fiona Stanley Hospital, Perth, WA; University of Newcastle, NSW

## Abstract

**Introduction:** Despite increased emphasis on education and training for patient safety in medical schools, there is little known about factors influencing decision making regarding patient safety behaviours. This study examined the nature and magnitude of factors which may influence opinions around patient safety related behaviours as a means of providing insights into how Australian doctors and medical students view these issues relative to members of the public.

**Methods:** A national, multicentre, prospective, on-line cross sectional survey was conducted using responses to hypothetical clinical scenarios. Three cohorts were surveyed - Australian enrolled medical students, medical doctors and members of the public.

Participant responses were compared for the different contextual variables within the scenarios and the participants’ demographic characteristics – student, doctor, member of the public, gender and age (if public or doctors)/ seniority in the course (if a medical student).

**Results:** In total there were 2602 medical student participants, 809 doctors and 503 members of the Australian public. Medical doctors were more likely than other cohorts to have statistically significant differences in how they viewed the acceptability of patient safety related behaviours; doctors were more tolerant of medical students not reporting concerning behaviours. Medical students’ opinions frequently demonstrated a ‘transition effect’, bridging between the doctors and publics’ attitudes, consistent with professional identity formation.

**Conclusions:** Opinions on the acceptability of medical students’ patient safety related behaviours were influenced by the demographics of the cohort and the contextual complexity of the scenario. Although the survey used hypothetical scenarios, doctors and medical students’ opinions appear to be influenced by cognitive dissonances, biases and heuristics which may negatively affect patient safety.

*‘Opinion is the medium between knowledge and ignorance’* Plato

## Introduction

Healthcare is increasingly viewed as a safety-critical industry ^1^. The importance of education and training for patient safety is now recognised by medical schools ^2-4^. Despite the increased emphasis on patient safety in health care, there are relatively few studies which examine the factors influencing health professionals’ opinions on decision making around patient safety issues ^5-15^. This is despite the understanding that patient safety is rarely just a technical issue, and is embedded in culture, organizational and professional politics ^16^.

The theory of planned behaviour (TPB) has been used as a framework to evaluate medical professionalism and patient safety related behaviours ^5 6 11 17-19^. The TPB is a useful model for explaining behaviours that are under volitional control. It specifies that intentions are the precursors of behaviours; the stronger the intention, the more likely the behaviour will be performed ^20^. Intention is influenced by three variables- the attitudes of the protagonist towards the behaviour, their perceptions of the social norms, and their perceived ability to perform the behaviour. Studies examining TPB and patient safety show that there have to be significant risks to patient safety before doctors raise concerns ^5^. Also different health professional groups (doctors, nurses, and allied health) have been shown to have unique behavioural factors which influence their intention to engage in patient safety behaviours ^10-12^, with doctors, particularly junior clinicians, most influenced by professional peer behaviour, i.e. their colleagues’ patient safety behaviours ^11^.

Research on factors that may influence incident reporting and ‘speaking up for safety’ demonstrate significant differences between patients and clinicians ^12 21^. Patients are more likely to report incidents associated with emotional or psychological harm ^21^, with the majority of patients’ defined incidents not considered to be patient safety incidents when reviewed by clinicians ^22^. There is little published literature on medical students’ opinions on acceptable professional behaviours related to patient safety issues and how these compare with qualified doctors and members of the public ^23 24^

The Professionalism of Medical Students study (PoMS) was designed as a body of research to examine which factors influenced opinions on medical students’ behaviours over a wide range of professionalism scenarios. The research sought to explore the effects of different contexts on professional decision making and assess whether demographic factors influenced opinions on the acceptability of behaviours. Although the scope of the PoMS work was to analyse opinions on a wide range of professionalism dilemmas, a number of these covered patient safety related topics such as fabricating results, infection controls, use of personal protective equipment, escalating concerns or reporting errors and the influence of hierarchy and health professional roles.

The aims of this study were to examine the nature and magnitude of factors which may influence opinions around medical student patient safety related behaviours as a means of providing insights into how Australian doctors and medical students view these issues relative to members of the public.

## Methods

### Design, ethics, setting and participants

This study was a national, multicentre, prospective, cross sectional survey. Ethical approval was obtained from UWA for each of the three recruitment cohorts: Australian enrolled medical students (HREC RA/4/1/8014), Australian medical doctors (HREC RA/4/1/9195), and Australian public (HREC RA/4/1/9278). We used a convenience sampling approach to survey medical students in Australia and New Zealand, members of the Australian public and Australian medical profession.

The methods and results of the medical student data, and validation of the survey instrument have been previously described in the *Medical Students’ opinions on professional behaviours* paper, referred to henceforth as the PoMS-I study ^17^. As the student only data analysis indicated that national factors had an effect on opinions about students’ professional behaviours ^17^, for this paper we limited our study to Australian populations.

In order to optimise recruiting a wide variety of Australian doctors and members of the public to participate in the study we utilised a range of approaches via conventional and social media. A social media webpage (Facebook Inc., Menlo Park, California) advertising the research project and link to the surveys was produced. To help notify medical doctors about the research project, the Australian Medical Association published information about the survey in their national news publication (Australian Medicine), and distributed it through their GP News network. The Australian Confederation of Postgraduate Medical Education Councils sent the survey information to their listed directors of postgraduate medical education for dissemination through their networks.

For the public, in addition to the social media advertising, the Australian Consumers’ Health Forum placed information about the research project and survey on their website. The anonymous, online surveys were closed when recruitment had plateaued (less than 10 responses/month for 3 months) in April 2018.

### Survey instrument

The surveys used in the PoMS research covered a wide variety of professionally challenging situations pertinent to medical students ^17^. Five of the scenarios related to a patient safety issue. Each scenario had two versions (vignettes) which contained a contextual variable identified *a priori* using the modified Delphi approach ^17^. For example, the protagonist in the scenarios could be either a junior or senior student, male or female, or another health professional. Likewise the context of the scenario could alter from agreeing/not agreeing to perform a procedure or attending/not attending a clinical placement. The online survey (SurveyMonkey Inc., San Mateo, California) was designed such that each participant received a random version of each scenario to allow comparison of the contextual variables. This provided a means of analysing whether respondents were influenced by unconscious bias; that is, whether the seniority, gender or type of health professional involved influenced the respondents’ opinions on the acceptability of the behaviour.

The scenarios were constructed to encourage participants to reflect on the acceptability of behaviours which ranged from serious professionalism breaches to positive examples of professional behaviour. All scenarios ended with the sentence ‘*This student’s behaviour is…*’. Respondents gave their opinions on ‘acceptability’ using a four point Likert scale, with no option for equipoise. The survey is included in Appendix A; survey scenarios relating to patient safety themes along with their contextual variables are described in Table 1.

**Table 1.**
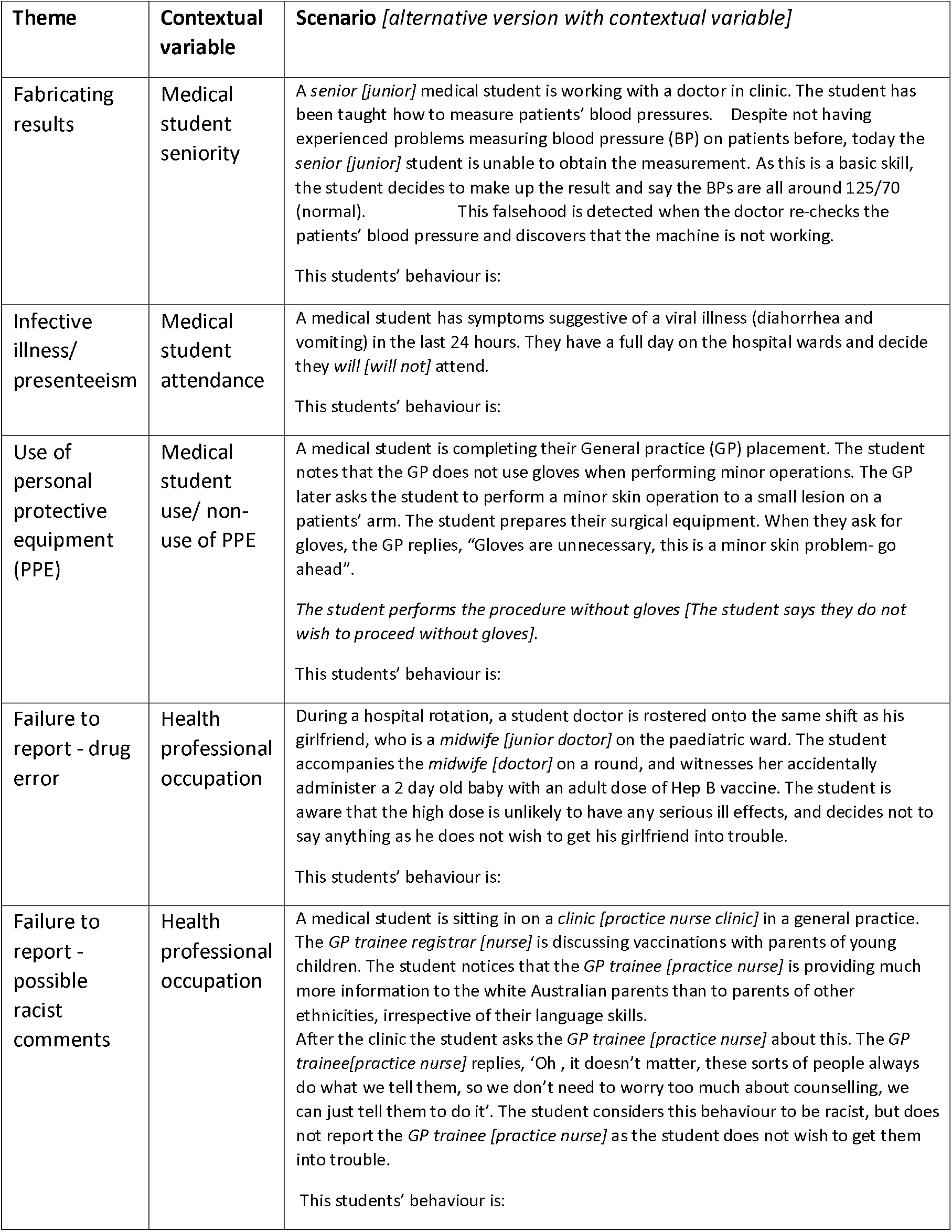
PoMS survey scenarios relating to patient safety themes (including contextual variables)

### Data and statistical analysis

Likert scale responses were combined into binary categories of all ‘acceptable’ responses against all ‘unacceptable’ responses and summarized using frequency distributions. The Chi-square or Fisher exact test was used to compare responses for the different contextual variables within the scenarios and the participants’ demographic characteristics – student, doctor, member of the public, gender and age (if public or doctors)/ seniority in the course (if a medical student). Null and blank responses were excluded for all questions, as were ‘prefer not to say’ responses for gender. SPSS Statistics 24 (IBM Corp) was used for data analysis and a *p*-value of <0.05 was considered statistically significant.

Figures in the Tables provided may not add up to 100 per cent due to rounding (nearest decimal place) and totals may vary due to some participants not completing all of the questions or for example, using option ‘prefer not to specify’ for gender question.

## Results

The demographic details of the survey respondents are presented in Table 2. The participation of all Australian medical schools resulted in larger numbers of student respondents (n=2602) compared to doctors (n=809) or the public (n=503). There were significantly more female participants in all three cohorts (p<0.001). We grouped doctors and members of the public according to age; public respondents were significantly older than the doctors who participated in the survey (p<0.001). Medical students were classified as ‘junior’ if they were in the first two years of a post-graduate entry course or in years 1–3 for an undergraduate course; by default, senior students were in the latter years of these courses. By performing subgroup statistical analysis for age and gender we were able to correct for the preponderance of female participants overall, and older public respondents.

**Table 2.** Demographic details of PoMS survey participants: Gender and Age/Seniority^⍰^.

| **Demographic factor: Gender (n)** | **Gender numbers (%)** |  |
| --- | --- | --- |
| **Member of the public (503)** | Male = 147 (29.2) | Female = 349 (69.4) |
| **Medical doctors (809)** | Male = 200 (24.7) | Female = 599 (74) |
| **Medical students (2602)** | Male = 1105 (42.8) | Female = 1439 (55.8) |

| **Demographic factor: Age/Seniority (n)** | **Age stratification numbers (%)** |  |  |  |  |
| --- | --- | --- | --- | --- | --- |
|  | Age <35 | Age 36-45 | Age 46-55 | Age 56-65 | Age >65 |
| **Member of the public (503)** | 164 (32.8) | 86 (17.2) | 93 (18.6) | 98 (19.6) | 59 (11.8) |
| **Medical doctors (809)** | 390 (48.4) | 242 (30) | 98 (12.2) | 57 (7.1) | 19 (2.4) |
| **Medical students<sup>Ⓜ</sup> (2602)** | 'Junior' 1214 (50.6) |  | 'Senior' 1184 (49.4) |  |  |

<sup>Ⓜ</sup> Medical students were classified as 'junior' if they were in the first two years of a post-graduate entry course or in years 1–3 for an undergraduate course; by default, senior students were in the latter years of these courses.

### Influence of participant demographics

Tables 3a and 3b illustrate how different participant demographic factors influenced the opinions expressed for the various scenarios. The demographic group that most frequently differed in their opinions on the acceptability of medical student patient safety related behaviours when compared to other groups were medical doctors (Table 3a).

**Table 3a.** Effect of participants’ demographic background on their responses to the PoMS patient safety scenarios.

| Scenario | Contextual variable | No. of public respondents stating either 'acceptable' or 'mostly acceptable' opinions on protagonist's behaviour/ No. of public respondents (%) | No. of student respondents stating either 'acceptable' or 'mostly acceptable' opinions on protagonist's behaviour/ No. of student respondents (%) | No. of doctor respondents stating either 'acceptable' or 'mostly acceptable' opinions on protagonist's behaviour/ No. of doctor respondents (%) | Comment on statistical significance<sup>a</sup> and if transition effect<sup>b</sup> |
| --- | --- | --- | --- | --- | --- |
| Fabricating results | Senior student | 9/232<br>(3.9) | 8/1184<br>(0.7) | 2/364<br>(0.5) | Public significantly more likely to consider it acceptable to fabricate results compared to student or doctor respondents; p=0.001 (students), p = 0.04 (doctors). |
|  | Junior student | 10/247<br>(4.0) | 8/1214<br>(0.7) | 3/396<br>(0.8) | Public significantly more likely to consider it acceptable to fabricate results compared to student or doctor respondents; p<0.001 (students), p = 0.007 (doctors). |
| Infective illness | Student attends | 14/253<br>(5.5) | 144/1208<br>(11.9) | 66/395<br>(16.7) | Doctors more likely to consider it acceptable to attend compared to student or public respondents.<br>Doctors compared to public, p<0.001;<br>Doctors compared to students, p= 0.002;<br>Students compared to public, p <0.001.<br>Transition effect. |
|  | Student does not attend | 201/214<br>(93.9) | 1141/1161<br>(98.3) | 348/348<br>(100) | Doctors more likely to consider it acceptable NOT to attend compared to student or public respondents.<br>Doctors compared to public, p<0.001;<br>Doctors compared to students, p= 0.022;<br>Students compared to public, p <0.001.<br>Transition effect. |
| Personal protective equipment (PPE) | Agrees to proceed without PPE | 45/235<br>(19.1) | 181/1156<br>(15.7) | 87/394<br>(22.1) | Doctors had highest rates for acceptability of student proceeding without using appropriate PPE, but no significant differences between demographic groups. |
|  | Refuses to proceed without PPE | 198/218<br>(90.8) | 1157/1180<br>(98.1) | 349/351<br>(99.4) | Doctors had highest rates for acceptability of student refusing to proceed without appropriate PPE.<br>Doctors compared to public, p<0.001;<br>Doctors compared to students, p = N/S;<br>Students compared to public, p <0.001.<br>Transition effect. |
| Failure to report- drug error | Midwife | 6/199<br>(3.0) | 71/1126<br>(6.3) | 14/349<br>(4.0) | No significant differences between demographic groups. |
|  | Junior doctor | 16/246<br>(6.5) | 66/1114<br>(5.9) | 39/372<br>(10.5) | Doctors had highest rates for acceptability of student not reporting a drug error by the junior doctor.<br>Doctors compared to public, p= N/S;<br>Doctors compared to students, p = 0.003;<br>Students compared to public, p= N/S. |
| Failure to report- racist comments | Practice nurse | 40/215<br>(18.6) | 322/1121<br>(28.7) | 111/354<br>(31.4) | Doctors more likely to consider it acceptable to 'not to report' practice nurse.<br>Doctors compared to public, p= 0.001;<br>Doctors compared to students, p = N/S;<br>Students compared to public, p= 0.002.<br>Transition effect. |
|  | Junior doctor | 38/229<br>(16.6) | 361/1094<br>(33) | 148/356<br>(41.6) | Doctors more likely to consider it acceptable 'not to report' junior doctor.<br>Doctors compared to public, p< 0.001;<br>Doctors compared to students, p = 0.003;<br>Students compared to public, p< 0.001.<br>Transition effect. |

<sup>a</sup> p-value calculated for combined 'acceptable' opinions for the different versions (vignettes) in each scenario. Chi-squared used for binary variables (Combined Acceptable/Mostly Acceptable vs. Combined Unacceptable/Mostly Unacceptable responses) unless category value <5 in which case Fishers' Exact test performed.
<sup>b</sup> Transition effect- noted if there was a pattern of participant responses which demonstrated incremental change from public→ medical student→ doctors

**Table 3b.** Effect of participants’ sub-group demographic background on their responses to the PoMS patient safety scenarios.

| Scenario | Contextual variable | Public<br>[Prev Ratio]<sup>c</sup> |  | Students<br>[Prev Ratio]<sup>c</sup> |  | Doctors<br>[Prev Ratio]<sup>c</sup> |  | Comment on statistical significance<sup>a</sup> |
| --- | --- | --- | --- | --- | --- | --- | --- | --- |
|  |  | Gender<br>(effect if male) | Age<br>(effect if older) | Gender<br>(effect if male) | Age<br>(effect if senior student<sup>d</sup>) | Gender<br>(effect if male) | Age<br>(effect if older) |  |
| Fabricating results | Senior student | NS<sup>a</sup> | NS | NS | NS | NS | NS | Although public significantly more likely to consider it acceptable to fabricate results compared to student or doctor respondents, no significant differences in sub-group comparison for either gender or age/ seniority of respondent. |
|  | Junior student | NS | NS | NS | NS | NS | NS | As per Senior student comment. |
| Infective illness | Student attends | NS | NS | 14% vs 10.1%; [1.39] | 16.2% vs 6.7%; [2.42] | NS | NS | Student respondents were only cohort to demonstrate that the gender (p=0.038) or seniority (p<0.001) of the student respondent influenced opinions. |
|  | Student does not attend | NS | NS | NS | NS | NS | NS | No significant differences in sub-group comparison for either gender or age/seniority of respondent. |
| Personal protective equipment (PPE) | Agrees to proceed without PPE | 28 vs 14.6%; [1.92] | 14.2 vs 30.1%; [0.47] | 20.0 vs 12.2%; [1.64] | NS | NS | 18.1 vs 26.3%; [0.69] | Sub-group comparison for either gender or age/seniority of respondent shows:<br>- age effect with older doctor (p= 0.05) and public respondents (p= 0.004) being LESS likely to consider it acceptable if student performs procedure without PPE<br>- gender effect with student (p<0.001) and public respondents (p=0.014) being MORE likely to consider it acceptable if student performs procedure without PPE. |
|  | Refuses to proceed without PPE | NS | NS | NS | NS | NS | NS | No significant differences in sub-group comparison for either gender or age/seniority of respondent. |

| Scenario | Vignette comparator | Public [Prev Ratio]<sup>c</sup> |  | Australian Students [Prev Ratio]<sup>c</sup> |  | Doctors [Prev Ratio]<sup>c</sup> |  | Comment on statistical significance<sup>a</sup> |
| --- | --- | --- | --- | --- | --- | --- | --- | --- |
|  |  | Gender (effect if male) | Age (effect if older) | Gender (effect if male) | Age (effect if senior student<sup>d</sup>) | Gender (effect if male) | Age (effect if older) |  |
| Failure to report-drug error | Midwife | NS | NS | 7.9 vs 4.8%; [1.62] | NS | NS | NS | Sub-group comparison for either gender or age/seniority of respondent shows:<br>- gender effect only for male student respondents (p=0.045) who were MORE likely to consider it acceptable if student did NOT report a drug error by the midwife. |
|  | Junior doctor | NS | NS | NS | NS | 17.0 vs 8.4%; [2.03] | NS | Sub-group comparison for either gender or age/seniority of respondent shows:<br>- gender effect only for male doctor respondents (p=0.019) who were MORE likely to consider it acceptable if student did NOT report a drug error by the junior doctor. |
| Failure to report-racist comments | Practice nurse | 28.6 vs 15.3%; [1.87] | NS | NS | 35.5 vs 21.4%; [1.66] | NS | NS | Sub-group comparison for either gender or age/seniority of respondent shows:<br>- gender effect only for male public respondents (p=0.029) who were MORE likely to consider it acceptable if student did NOT report the practice nurse<br>- seniority effect only for student respondents (p<0.001) who were MORE likely to consider it acceptable if student did NOT report the practice nurse. |
|  | Junior doctor | NS | NS | NS | 42 vs 23.9%; [1.76] | 51.1 vs 38.7%; [1.32] | NS | Sub-group comparison for either gender or age/seniority of respondent shows:<br>- gender effect only for male doctor respondents (p=0.038) who were MORE likely to consider it acceptable if student did NOT report the junior doctor<br>- seniority effect only for student respondents (p<0.001) who were MORE likely to consider it acceptable if student did NOT report the junior doctor. |

<sup>a</sup> *p*-value calculated for combined 'acceptable' opinions for the different versions (vignettes) in each scenario. Chi-squared used for binary variables (Combined Acceptable/Mostly Acceptable vs. Combined Unacceptable/Mostly Unacceptable responses) unless category value <5 in which case Fishers' Exact test performed. NS used to denote 'not statistically significant'. i.e. $p \geq 0.05$
<sup>b</sup> Transition effect- noted if there was a pattern of participant responses which demonstrated incremental change from public→ medical student→ doctors
<sup>c</sup> [Prevalence ratio calculated for values of $p < 0.05$ ]; Prevalence ratio = [prevalence of respondents stating 'acceptable' in demographic group/prevalence of respondents stating 'acceptable' in comparison demographic group] (Thompson et al. 1998). For example, the vignette involving a student attending a clinical placement despite recent illness, male students ( $p = 0.038$ ) and senior students ( $p < 0.001$ ) were significantly more likely to consider this acceptable behaviour than female or junior students respectively; such that male and senior students were x1.39 and x2.42 more likely to respond that this behaviour was either 'acceptable' or 'mostly acceptable'.
<sup>d</sup> Senior students defined as either undergraduate entry years 4–6 or post-graduate entry years 3–4. Junior students defined as either undergraduate entry years 1–3 or post-graduate entry years 1–2.

Half of the scenarios demonstrated a ‘transition effect’ in which there was either a progressively increasing or decreasing level of acceptability for the behaviour described determined by whether the respondent was a member of the public, medical student or doctor (Table 3a). Transition effects were evident in the scenarios involving attendance at clinical settings with an infective illness, and those related to ‘failure to report’ situations.

### Influence of unconscious bias

To assess for possible effects of unconscious bias, each individual participant’s survey had a randomly allocated vignette for the scenarios to determine whether participants would be influenced by the contextual variable within that scenario.

The contextual variable for the fabricating results scenario was the seniority of the medical student protagonist. There were no significant differences in opinions for any demographic group (public/student/doctor; Table 3a) or sub-group (respondents’ gender/age/seniority; Table 3b).

Conversely the two ‘failure to report’ scenarios had the student protagonist interacting with people in differing health professional roles. Doctor participants were the only demographic group to have significantly different responses for the acceptability of ‘failure to report’ behaviour depending on the type of health professional involved for both ‘failure to report’ scenarios (Table 3a). Male doctor respondents were significantly more likely to consider it acceptable for medical students not to report junior doctors for either drug errors or potentially racist comments (Table 3b).

## Discussion

This is the first study to triangulate opinions on a national level from the public, medical students and qualified doctors on a range of patient safety scenarios involving medical students. The results provide a unique insight into the nature and magnitude of factors which influence opinions on patient safety and the challenges in this area for Australian health professionals.

The only other study to triangulate the public, students’ and doctors’ opinions on professional behaviours was performed at a single UK medical centre, and recruited the public by means of approaching people attending a paediatric outpatients ^23^. The total number of participants surveyed were 130 (54 in the public cohort). Although the scenarios used differed from the PoMS survey, a number of similar themes were covered such as fabricating results and inappropriate attendance at a clinical setting. Brockbank *et al* showed that the public, then doctors, and lastly students appeared to have incrementally increasing tolerance for professionally concerning behaviours by medical students and attributed this to the fact that the public inherently approach and judge professional behaviours from a different perspective to doctors and students ^23^.

Regarding possible limitations in study design, we used convenience sampling methodology. Selection bias was minimised by using a wide range of methods to promote recruitment and continuing the survey process until there were at least 500 participants in each cohort ^25^. We have no denominator data for the doctor and public participants, but PoMS-I calculated the medical student participation rate to be 15.2% of all Australian enrolled medical students, making it one of the largest medical student datasets in the literature ^17^.

A third-party perspective was used in the survey’s vignettes to minimise respondent’s social desirability biases ^26^. This format has also been used and endorsed in other studies ^24 27^. Although asking survey participants’ opinions on the protagonist’s behaviours could be considered too positivist/empirical an approach for the complex subject of patient safety, many psychological studies of human behaviour have used and validated similar approaches ^27^.

Human behaviour, decision making, and cognitive biases have been the subject of much research, with many questions still unanswered, particularly with respect to healthcare professional behaviours ^6-15 19 28^. To assist with discussing the results of this study and the implications for patient safety, we have provided a summary of key relevant theories and constructs related to human decision making and biases in Table 4.

**Table 4.**
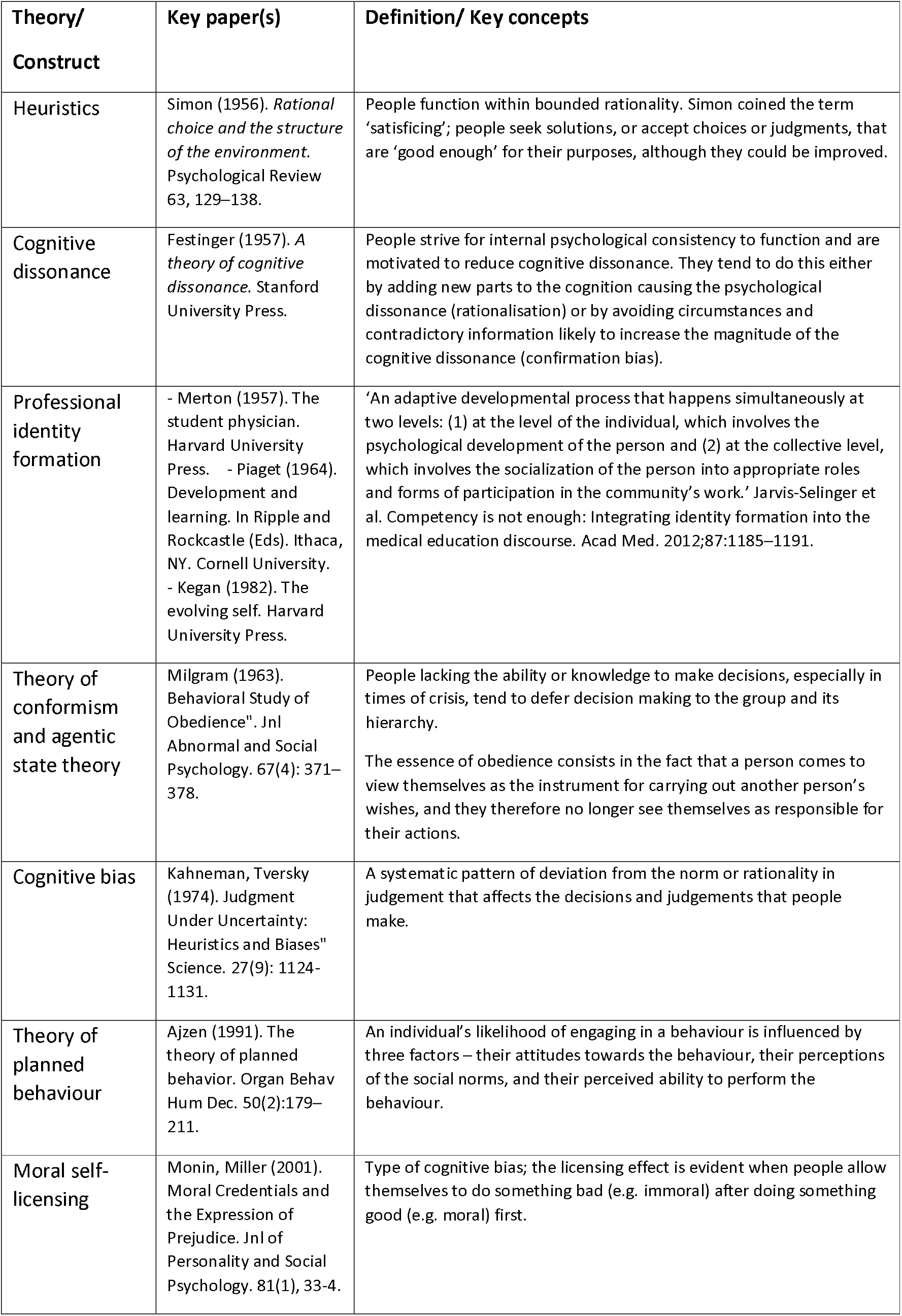
Major theories and constructs related to human decision making and biases.

The safety themes examined in this paper covered a wide range of clinical situations (Table 1). The first scenario related to a medical student fabricating the results of patients’ blood pressure readings. It is notable that there were very low levels of acceptability for this in any of the survey cohorts (public, students or doctors), no contextual effect (whether it involved a junior or senior student), and sub-group analysis for gender or age/seniority showed no evidence of unconscious bias. However public respondents were significantly more likely to consider it acceptable for the student protagonist to fabricate results compared to the medical student or doctor respondents. This implies that whilst the public appreciates the significance of fabricating results, they are less stringent in their expectations around this compared to doctors or students.

The next survey scenario related to infection control/presenteeism. The doctor respondents’ results were the cohort most likely to consider it acceptable behaviour for the medical student protagonist to either attend or not attend a clinical placement after a probable viral gastric illness, depending on which contextual variable they were presented with in the online survey.

Cognitive dissonance typically occurs when individuals experience two or more inconsistent beliefs, or when their behaviour is inconsistent with their beliefs/values ^7^ (Table 4). Doctor respondents as a group displayed cognitive dissonance in how they viewed the medical student’s behaviour in the infection control/presenteeism scenario. Although doctors were most likely to concur with the correct behaviour (when the student does not attend the clinical setting), when doctors were presented with the contextual alternative of the student attending, they were also the group most likely to consider this behaviour as acceptable. Doctors were more than three times more likely to consider it acceptable for a medical student to attend in these circumstances compared to members of the public. The PoMS data do not provide us with explanations for why this dissonance occurs and what heuristics may be operating, but doctors as a professional group are at high risk of presenteeism ^29 30^.

Medical student respondents who were male and in the latter stages of the course were more likely to consider that the behaviour of the recently unwell protagonist attending the clinical placement was acceptable, and hence more congruent with qualified doctors’ responses. This change in the medical students’ opinions as they progress through the course is a good example of the ‘transition effect’. Research on medical professionalism and sociology supports the concept and recognises the importance of professional identity formation (PIF, Table 4) ^31 32^. Merton in 1957 was one of the first to describe this phenomenon, when he noted medical education’s core function to *“shape the novice into the effective practitioner of medicine, to give him the best available knowledge and skills, and to provide him with a professional identity so that he comes to think, act, and feel like a physician”* ^33^.

PIF requires the individual to accept the norms of behaviour established by the community they wish to identify with ^34^. Some discretion is given for personal habits/beliefs, but the core attitudes and beliefs of the community are essentially nonnegotiable, resulting in either sanctions or exclusion from their community of practice ^34-36^. Each medical student’s journey is unique and will be influenced by their role models, mentors and experiential learning ^31-33 36-38^.

Medical students, like doctors, will be cognisant of the patient safety implications of attending a clinical area when recently unwell, yet more than 10% of student respondents stated that it was acceptable for the protagonist to attend in these circumstances. As previously noted, the study data do not provide us with explanations for why medical students’ opinions change as they progress through the course, but it may be a hidden curriculum effect ^32^ with students mirroring doctors’ presenteeism behaviours, or related to moral self-licensing (Table 4); by attending the placement, the student perceives that they are helping the clinical team, which mitigates the possible harm caused by attending.

These complexities in human decision making were also evident in the PPE scenario- the student protagonist either agreeing or refusing to perform a minor surgical procedure if their preceptor did not provide them with appropriate PPE. Incongruous opinions were expressed again by the doctor cohort. Doctors were the group most likely to accord with the correct behaviour (the student protagonist refusing to perform the procedure without appropriate PPE), but when they were presented with the contextual alternative of the student agreeing to perform the procedure without PPE, doctors were the group most likely to consider the behaviour as acceptable. Analogous to the infection control/presenteeism scenario, male medical students were significantly more likely to align with the doctors’ responses.

The possible factors influencing these cohorts’ opinions merit discussion as they provide useful insights into patient safety psychology. In the scenario, the student observes their GP preceptor not using gloves when operating, so one factor could be the student protagonist modelling their preceptor’s behaviour. Several studies have found strong relationships between observation and participation in terms of unprofessional behaviour; medical students were more likely to perceive unprofessional behaviours as being ‘acceptable’ if the student had either observed or participated in the activity ^39 40^. Students may try to normalise the professionalism breaches that they witness or participate in, a process which Monrouxe described as *‘habituation’* ^38^, or manage the dissonance by emotional trading- the benefits of gaining the clinical experience being offered outweigh the potential risks to the patient or student of not using PPE ^7^.

Another factor may relate to positional obedience; the protagonist’s actions may be justified as the instruction was given by an authority figure. Milgram’s classic studies ^41^ on the factors influencing obedience and conformism demonstrate that the perception of a legitimate authority is a powerful influencer on behaviour (Table 4). There are relatively few studies which specifically examine obedience behaviours in health care ^28 42-44^, but most support Milgram’s findings that people lowest in the hierarchy are most susceptible to following orders, despite the irrationality of the instructions ^41^. In our study, public respondents were the cohort least likely to consider it acceptable for the protagonist to refuse to follow their GP preceptor’s instructions. This implies that the public, the cohort which have been described in healthcare as suffering from epistemic injustice – injustice related to a lack of knowledge ^22^, may be most susceptible to the positional obedience effect.

The influence of the type of health professional involved in a professionalism dilemma and the context of what is ‘reportable’ were explored in the two ‘failure to report’ scenarios. The first scenario involved a drug error-an incorrect dose of vaccine is administered to a newborn, further complicated by the student protagonist being in a relationship with the health professional involved. Table 3a shows that whilst the public and medical students did not appear to factor in the type of health professional involved when making decisions about the acceptability of reporting drug errors, qualified doctors were significantly less likely (p<0.001) to consider it acceptable to report the health professional if the individual was a doctor.

Analysis of the results of doctors’ opinions in both failure to report scenarios indicates that doctors were the cohort most likely to consider it acceptable ‘not to report’ for both these scenarios. This suggests that doctors have a cognitive bias towards not reporting other doctors. Social psychologists describe the phenomenon of ‘in-group’ bias to describe the bias of individuals who identify as being part of a group to favour their group ^13^. Most studies examining the effects of implicit bias in health care have focused on implicit bias towards patients, but there is a growing literature on the factors which influence speaking up for patient safety and/or whistleblowing ^5 8-12 14 15 45^.

Blenkinsopp *et al’s* systematic review on ‘Whistleblowing over patient safety and care quality’ ^14^ noted that research in this area almost exclusively focussed on nurses rather than doctors (>80% of the studies) and attributed this to the emphasis in nursing on patient advocacy. Their paper documented the effect of organisational and occupational cultures on incident reporting, a phenomenon also described in Australian health care ^11 46^. ‘In-group’ bias may be the most deleterious effect of professional identity formation. Wakefield *et al* and Jorm both describe doctors as often being disconnected and at odds with the system they work in ^11 46^, with a medical culture *‘characterised by focussing on individual concepts of clinical work, clinical purism and opaque accountability’*, and junior doctors being ‘*quickly socialised to these values and behaviours’* ^11^.

The PoMS survey results validate the effects of occupational culture on opinions for patient safety behaviours ^5 8-12 21 28 38 44 45^. The scenario involving possible racist comments shows significant transition effects; the public, junior then senior medical students, and finally doctors demonstrate progressively increasing tolerance for not reporting a junior doctor in these circumstances. The overall result is that doctors are more than 2.5 times more likely to consider it acceptable not to report compared to public respondents. These findings resonate with O’Hara *et al’s* research on patient self-reporting of what they perceive to be reportable safety issues ^21^; patients were significantly more likely than clinicians to consider dignity and respect issues as being within the remit of patient safety reporting. In this PoMS scenario, the public appear to be more sensitive and less tolerant of the lack of respect shown to non-Caucasian patients by the health professional protagonists.

From a theory of planned behaviour (TPB) perspective ^6 19 20^, the individual’s attitudes, the subjective norms and perceived behaviour controls all influence behavioural intention. Hence it follows that the more ambiguous/contextually complex the dilemma, the more likely that there will be a multitude of factors affecting the three drivers of behavioural intention. The PoMS scenarios illustrate how increasing the contextual complexities related to decision making for professional dilemmas leads to an increased range for what participants considered to be ‘acceptable’ behaviours. For example, compare the relatively homogenous responses from the survey cohorts for the fabricating results scenario to the failure to report scenarios, which demonstrate significant differences between the cohorts and sub-group analysis (Table 3b).

Overall, this paper’s findings reveal several concerning results. Specifically, how leniently doctors viewed some of the patient safety related behaviours by medical students, the negative effects of professional identity formation, and possible cognitive biases which appear to influence opinions. Modern health care relies on functional teams and hierarchies, and significant medical errors tend to occur in complex environments ^47^. The Australian Commission on Safety and Quality in Health Care defines safety culture as *‘a product of individual and group values, attitudes, perceptions, competencies and patterns of behaviour that determine the commitment to, and the style and proficiency of an organisation’s health and safety management’* ^48^. This paper illustrates the complexities of patient safety-that it is intrinsically linked to organisational and professional politics ^16^ with often significant divergences in how the public and doctors view these issues. Health care providers and educators need to factor in the cognitive biases that exist within the individual and groups as a means of managing the complexity of personal beliefs, professional norms and work place cultures to improve patient safety ^6-15 47^.

## Conclusions

This research demonstrates that opinions on the acceptability of behaviours related to patient safety themes by medical students are influenced by the demographics of the participant, and the contextual complexity of the scenario. The Australian doctors, and to a lesser degree, medical students who were surveyed, had differing opinions regarding medical student patient safety related behaviours than members of the public. This was most apparent when related to infection control issues such as attending health care settings when recently unwell, refusing to comply with instructions, or reporting concerns or errors that involved a doctor. Medical students’ opinions on professional behaviours often bridged the attitudes of the Australian public and doctors, adding further support to the concept of medical student professional identity formation ^31-33. 4^The GMC states that *‘Professionalism is not about doing the minimum – it is about doing what is necessary to protect patients’*. Although this study was directed towards hypothetical medical student behaviours, our results indicate that the cognitive dissonances, biases and heuristics which appear to influence doctor’s opinions on medical students’ behaviours may present significant challenges to patient safety in clinical practice. We suggest that efforts to improve patient safety should recognise these factors and consider how best to address them to ensure that patient care is protected.

## Data Availability

Not available on other sites at present

## Notes

### Competing Interest Statement

The authors have declared no competing interest.

### Clinical Trial

Australian enrolled medical students (HREC RA/4/1/8014), Australian medical doctors (HREC RA/4/1/9195), and Australian public (HREC RA/4/1/9278).

### Funding Statement

No funding

### Author Declarations

Ethical approval was obtained from UWA for each of the three recruitment cohorts: Australian enrolled medical students (HREC RA/4/1/8014), Australian medical doctors (HREC RA/4/1/9195), and Australian public (HREC RA/4/1/9278).

